# Caregiving for People with Lewy Body Dementia During A Pandemic: A Qualitative Analysis of Caregivers who Completed Resilience Coaching

**DOI:** 10.1101/2025.07.17.25331719

**Authors:** Arden Chan, Ejew Kim, Madeleine E. Hackney

## Abstract

**Background:** The Activity Theory of Aging suggests sustained activity aids older adults to maintain high quality of life. Caregiving resilience coaching like the Dreaming Together (DTog) program may have helped caregivers for people with Lewy body dementia (LBD) sustain supportive activity throughout challenges from caregiving and pandemics.

**Objectives:** To determine experiences and attitudes on sustaining activities during the COVID-19 pandemic of LBD caregivers participating in caregiving resilience coaching two years into the pandemic.

**Methods:** 13 participants (10 female; age=67.7 (9.9) years; Caregiver Quality of Life score=36 (12.9), “good”; Zarit Burden Interview score=72 (18.8), “mild to moderate”) were recruited to participate in an Exit Interview held over Zoom right after DTog completion, between February and August of 2022. Participant responses were coded into themes through NVivo 12 software, for analysis relative to the Activity Theory of Aging.

**Results:** Main themes: 1) Technology: An imperfect solution to social isolation; 2) Self-care is a necessity even during a crisis; 3) Caregivers Report A Reinforcing Negative Cycle of Declining Care Recipient Health and Adaptability.

**Conclusion:** LBD caregivers, after completing the DTog resilience coaching program, maintained self-care activities that improved life satisfaction. Programs and studies promoting resilience-building activity should be considered to supporting LBD caregiving.

## Introduction

### Lewy Body Dementia: Symptoms and Treatment

Lewy Body Dementia (LBD) is an umbrella diagnosis for degenerative dementia-related diseases in older adults, including Dementia with Lewy Bodies (DLB) and Parkinson’s Disease Dementia (PDD/PD). LBD is the second most common cause of dementia after Alzheimer’s Disease (AD), affecting approximately 1.4 million Americans (Galvin, 2019). However, LBD may have greater impacts on daily life than AD due to increased motor-cognitive dysfunctions, sleep disorders, behavioral and emotional changes, and autonomic symptoms (i.e., postural dizziness, fatigue, mucosal dryness). Furthermore, LBD patients have a greater risk of hospitalization and mortality than AD patients due to more rapid disease progression and functional decline (Capouch, Farlow, & Brosch, 2018; Galvin et al., 2010).

Those afflicted with DLB have both cognitive and motor impairments at onset, whereas those with PDD symptoms tend to first exhibit motor impairments and then develop cognitive impairments over time (Gomperts, 2016). For AD, some prescriptions can alleviate dementia and its symptoms, such as memantine and cholinesterase inhibitors––rivastigmine, donepezil, and galantamine; however, these treatments have side effects and are mainly effective for dementia in the mild to moderate stages (Ellis, 2005). Several of these medications approved for AD are also used in treating DLB and PDD, but with varying success (Capouch et al., 2018). PDD-specific medications like carbidopa-levodopa are popularly used to manage some motor symptoms like tremor and rigidity, but have adverse effects including hallucinations, sleep disturbance, confusion, and motor complications with increased fluctuations and dyskinesia due to growing “OFF” time with extended drug use, which also deteriorates quality of life (Jankovic & Tan, 2020; Gandhi & Saadabadi, 2023).

Health literacy, language, socio-cultural, financial, and other barriers prevent access to effective treatments and healthcare. Dementia progression may be exacerbated by comorbidities, where it becomes difficult for patients to self-manage their condition (Chen et al., 2019).

### Lewy Body Dementia Caregivers

Caregivers play a critical role in the medical community as non-healthcare workers by looking after care recipients outside of hospital care. However, LBD patients’ perpetual needs increase the difficulty and costs of LBD caregiving. Out of the many subtypes of dementia, LBD diagnosis has the larger burden due to higher medical costs and lengthier hospital visits, which is nearly twice that of AD, the most prevalent subtype (Chen et al., 2019; LBDA, 2022). The conditions requiring these higher costs consists of falls (21.3%), urinary incontinence or infection (15.2%), psychiatric conditions (depression 4.9%, anxiety 3.4%), dehydration (4.2%), delirium (3.3%), orthostasis (2.7%), sleep disorders (1.9%), hallucination (1.2%), and delusion (<1%) (Chen et al., 2019). If caregivers can address, prevent, or manage these symptoms at home, then there is potential to reduce the costs of LBD (Chen et al., 2019). However, caregiver burden can include increased stress, anxiety, and decreased social interactions, which ultimately affect their personal health and lifestyle (Galvin et al., 2010). Caregivers may face depression and anticipatory grief, identity and existential crisis, and discrepancies in emotional and physical support from a spouse with dementia (Baikie, 2002; O’Shaughnessy, Lee, & Lintern, 2010).

Caregivers are also prone to caregiving burnout. Despite these issues, current modern medicine and programs tend to focus on the treatment for those directly affected by the diseases and provide little support to the caregivers.

While little is known about LBD caregiving, previous studies have investigated the stress and burdens related to AD caregiving. For instance, AD caregivers are susceptible to feelings of social isolation and depression with greater risks of medical illnesses and feelings of guilt (Galvin et al., 2010). Support interventions were also identified, such as social support networks, supportive familial and peer relations, and physician education (Galvin et al., 2010). However, AD caregiving strategies and interventions may not be applicable to LBD caregiving as LBD progresses more rapidly than AD, thus the loss of independence occurs earlier in LBD and causes greater dependency on the caregiver (Galvin et al., 2010).

Currently, few studies and programs support caregivers for people with LBD. To help people with LBD adjust to their new life with dementia, it is essential to acknowledge the personal wellbeing of caregivers (Bhimani, 2014). Caregiving especially needs attention for LBD as two-thirds of dementia caregivers are women, and thirty percent of caregivers are 65 years or older (Alzheimer’s Association, 2022; University of Michigan, 2017). When the caregiver is an older adult, caregiving difficulty and caregiver burden increase because older adults are often excluded in research and lack proper education on caregiving (Petrovsky et al., 2022; Taylor et al., 2012). In fact, research has shown that programs including older adults in research and educating them on disease and health advocacy help adults navigate through their challenges and realize that they are not alone in their health journeys (Hart et al., 2017).

### COVID-19 and Caregiving

While caregiving was already demanding before, caregiver burden may have significantly increased due to the coronavirus-19 (COVID-19) pandemic. COVID-19––an infectious respiratory disease caused by the SARS-CoV-2––presented numerous challenges to daily life, including a shortage of daily necessities, nationwide panic, and delays in public services, especially in the medical field. Significantly, social activities were heavily restricted due to the enforcement of social distancing. While 81% of those who passed away due to COVID-19 in 2020 were 65 years old or older (Tejada-Vera & Kramarow, 2022), caregivers experienced greater difficulty accessing the already limited resources and programs to caregiving support. With fewer caregiver resources and avenues of support overall, it is unknown how LBD caregiving and caregiver burden have been impacted by COVID-19 and the extent of the consequential impediments to caregiving.

### Activity Theory of Aging

Our investigation of participant experiences and attitudes regarding caregiving during COVID-19 is grounded on the *Activity Theory of Aging*, a theory guided by a Functionalist perspective that the social equilibrium experienced by middle aged adults should continue into older age years for benefits to themselves and society (Griffiths & Keirns, 2015; Havighurst, 1961). As young adults reach their prime, they establish a set of routines and activities that satisfy their psychosocial needs, which brings personal happiness and life satisfaction. To sustain happiness in older adults, these needs must remain satisfied throughout the aging process.

Therefore, the Activity Theory focuses on the positive relationship between the level of activity participation and life satisfaction in older adults, where the more active and involved an older adult person is, the greater the degree of happiness they will experience (Griffiths & Keirns, 2015; Havighurst, 1961; Knapp 1977). Consequently, activities are important to the needs of older adults as well. However, biological changes, social norms, and economic factors may prevent older adults from maintaining the same level of activity and engagement. Thus, to satisfy the same psychosocial needs and adapt to the changes that come with age, older adults need to find substitutes for the activities which they must give up (Havighurst, 1961). However, caregiving and COVID-19 became new variables that obstructed older adults from engaging in their activities.

### DREAMing Together (DTog): A Caregiver Empowerment Program

This study reports on the use of the DREAMing Together (DTog) program to investigate the extent to which caregiving was influenced by COVID-19 and the community response to the viral pandemic within six months of the pandemic onset. The methods of DTog derive from our prior program, Developing Research Education and Advocacy for Multicultural Seniors (DREAMS), which was designed to enhance research involvement and advocacy within older adults (Bay et al., 2020; Bay et al., 2023; Hart et al., 2017; Perkins et al., 2019; Schindler et al., 2022; Shah et al., 2023a; Shah et al., 2023b). DREAMS was reiterated for DTog specifically to supply education and coaching to caregivers for people with LBD and related dementias. See **Supplemental 1** for more information.

#### Purpose

Based on the Activity Theory of Aging which posits that a more active lifestyle is associated with higher levels of life satisfaction in older adults, we hypothesized that sustained activity through the pandemic would allow caregivers to be resilient. We aimed to understand the thought processes, actions, attitudes, beliefs, perspectives, and experiences of LBD caregivers who underwent DTog, a caregiving resilience coaching program during COVID-19. We administered an exit interview and used the Activity Theory of Aging to guide our analysis of the responses of caregiver participants, via recordings and transcripts, who completed the 8-week DTog program,.

## Methods

The Emory University institutional review board reviewed and approved this protocol (IRB00080676). All participants provided informed consent before participating. The DTog program began in September 2021. The COVID-19 impact data were collected in Exit Interviews from February 2022 to August 2022.

### Recruitment

Caregivers for people with DLB and PDD in and near the Atlanta metro area were recruited for DTog. We recruited 21 adults from the Emory LBD Roybal Center of Excellence registry and LBD or PD support groups. Participants were also recruited by word of mouth, from other study registries, and through foundation newsletters. Inclusion criteria for participants were that they could read, speak, and understand English, were 40 years and older, a family care partner of a spouse or life partner with LBD. Potential participants received a phone call to determine eligibility, discuss the DTog program, and schedule further meetings. After consent at the baseline visit, the DTog binder with the curriculum was sent to the participant via email or mail. Participants were compensated for participation in the assessments.

### The COVID-19 Caregiving Exit Questionnaire Interviews

Motivated by previous studies and questionnaires on the health of older adults during COVID-19 (Cawthorn et al., 2020; Cohen et al., 2020; Harkness 2020), the team developed a COVID-19 Exit Questionnaire to be administered by interview at the end of the of DTog program. To develop this questionnaire, the team brainstormed questions by considering the challenges of living in COVID-19 from personal experiences and observation of others’ experiences primarily from DTog participants. After generating a preliminary list of questions, the team narrowed this list down to six questions that prompted participants to reflect on their caregiving experiences during COVID-19, with respect to their health, care recipient health, and the caregiving experience during COVID-19.

At the Exit interview, participants were first asked questions related to their quality of caregiving during COVID-19 and were probed to share personal experiences. Next, coaches asked questions pertaining to the participants’ changes to personal life, including social and family activity (**Table 1)**. Staff encouraged participants to describe the challenges encountered during caregiving and skills that participants would have liked to develop to overcome these challenges. Exit interviews lasted on average 25 minutes, ranging from 15 to 45 minutes.

**Table 1.** DREAMing Together COVID-19 Exit Questionnaire, Assessment Interview.

|  |  |
| --- | --- |
| **Question** | **Sample Probes** |
| 1. How has your caregiving been impacted by the pandemic? | • What has caregiving been like due to the pandemic? Have you been doing a lot? |
| 2. What habits have changed? What routines have changed? What friendships or relationships have changed and how? | • What did your social activity/day-to-day schedule look like? |
| 3. How has the pandemic affected your partner's condition? | Was there anything that changed for your partner because of the pandemic? (Exercise routines, daily activities, etc.) |
| 4. How has the pandemic altered the way you use healthcare? | What was healthcare like for you during COVID-19? Has there been any changes to accessing healthcare? |
| 5. What are some challenges/struggles you are currently encountering? What are some ways you have dealt with new obstacles during COVID? | How did you adjust to the changes that occurred because of COVID-19?<br>What became easier? What became more difficult? |
| 6. What skills or aspects of life would you like to develop/improve during COVID? | Was there anything—skills, abilities, access to information—that could have made your life easier? |

### Quantitative Measures

After informed consent, DTog participants were administered a Project Health Demographic survey to record demographics and medical questions including race/ethnicity, age, gender, medical conditions, etc. Caregivers were also asked “in general, how would you rate the quality of your life?” on a 100-point scale (with 100 correlating to a very high quality of life).

The Zarit Burden Interview (ZBI) was used to measure caregiver burden on dementia caregivers. ZBI is an 88-point scale questionnaire that combines 22 questions each rated on a 5-point Likert scale. For each question, caregivers indicate the frequency of their feelings with the following scale: never (0), rarely (1), sometimes (2), quite frequently (3), or nearly always (4). A score of 0 to 21 indicates little or no burden, 21 to 40 indicates mild to moderate burden, 41 to 60 indicates moderate to severe burden, and 61 to 88 indicates severe burden.

### Thematic Data Analysis

Exit interviews were transcribed by Otter AI and later reviewed and refined for accuracy by research assistants. NVivo 12 software was used to facilitate all data organization and analysis. One primary and 3 secondary coders created a codebook of general and recurring themes on caregiving from participant response: A preliminary codebook was developed after reading the transcripts, then as more information and patterns were identified, recurring words and phrases from the raw data were coded and sorted under subthemes. All coders independently coded the transcript, and the primary coder reviewed and reconciled the coding differences with agreement by secondary coders and additional review by PI. The participants in this paper are referred to by an identifier assigned to them through a unique alpha-numeric code using the letter “C” followed by a number, e.g., “C3”. These identifiers do not correlate to the real identifiers used in the study.

The Activity Theory of Aging guided the thematic analysis of qualitative data, resulting in several important themes. Information and responses were categorized into three main themes: the DTog-educated caregivers’ response to COVID-19 obstacles regarding the act of caregiving, themselves, and the care recipient. Each subtheme identified successful and unsuccessful adaptive behaviors against COVID-19 (See **Table 2**).

**Table 2.** Thematic analysis of DTog Exit Interview.

| **Table 2.** Thematic analysis of DTog Exit Interview |  |
| --- | --- |
| **Main Theme** | **Subthemes** |
| Technology: An Imperfect Solution to Social Isolation | • Using and adapting to use technology helped caregivers to tackle some COVID-inflicted restrictions to daily, healthcare, and social activities<br>• Unresolved socioemotional disconnection and communication issues due to overpowering impact of physical isolation<br>○ Lack in access and benefit of support groups exacerbated despite technological adaptation |
| Self-Care is A Necessity Even During A Crisis | • The dominating and continuous nature of LBD caregiving during COVID-19 led to deterioration of caregiver mental, emotional, and physical well-being and risk of burnout<br>○ Reduction in social and physical activity due to pandemic and caregiving restrictions heightened emotional burden<br>• Social support and self-focused activities alleviated mental and emotional caregiving burden<br>• Adoption of positive mindsets allowed resilience and adaptive ability in caregivers |
| Caregivers Report A Reinforcing Negative Cycle of Declining Care Recipient Health and Adaptability | • Caregivers attribute care recipient functional decline to social isolation<br>• Caregivers highlight inadaptability in care recipients due to functional decline, which further hinders activity and functional improvement |
|  | Caregivers note socialization to support care recipient emotionally and cognitively |

## Results

### Participant Demographics

Of the 21 recruited participants for the DTog program, eight who were unable to participate in the Exit Interview were excluded: Six dropped out of the program before starting the modules, one finished the modules but did not complete the interview, and one had to postpone their learning beyond August of 2022. We present demographic and clinical characteristic data from 13 participants who completed the DTog modules and participated in the Exit Interview.

The participants’ average age was M = 67.7 years (SD = 9.9) and ranged from ages 53 to 83 years. Ten were women and three were men. Eight were White/Caucasian, one was Asian, one Hispanic or Latino, two Black/African American, and one multiracial. Participants had M = 16.7 (SD = 1.3) years of education, which is equivalent to a bachelor’s degree. Additionally, participants scored M = 36 (SD = 12.9) on the ZBI, indicating mild to moderate burden. Overall, participants reported having a good quality of life on the Quality-of-Life scale, M = 72 (SD = 18.8) between a range of 43 to 100 (**Table 3**).

**Table 3.** COVID-19 Exit Interview DTog Participant Demographics.

| **Table 3. COVID-19 Exit Interview DTog Participant Demographics** |  |
| --- | --- |
|  | N (%) / Mean (SD)<br>n=13 |
| Age (y) | 67.7 (9.9) |
| Gender |  |
| Male | 3 (23%) |
| Female | 10 (77%) |
| Education (y) | 16.7 (1.3) |
| Race |  |
| White/Caucasian | 8 (61%) |
| Black/African American | 2 (15%) |
| Asian | 1 (8%) |
| Hispanic/Latino | 1 (8%) |
| Multiracial | 1 (8%) |
| ZBI Score (/88) | 36.0 (12.9) |
| Quality of Life (/100) | 72 (18.8) |
| Marital Status |  |
| Single | 0 (0%) |
| Married | 13 (100%) |
| Separated/Divorced | 0 (0%) |
| Widowed | 0 (0%) |
| Veteran |  |
| Yes | 1 (7.7%) |
| No | 12 (92.3%) |
| Housing |  |
| House/apartment/condominium | 12 (92.3%) |
| Senior housing (independent) | 1 (7.7%) |
| Assisted living | 0 (0%) |
| Relative's home | 0 (0%) |
| Other | 0 (0%) |
| English first language |  |
| Yes | 12 (92.3%) |
| No | 1 (7.7%) |
| Transportation |  |
| Drive own vehicle | 13 (100%) |
| Family/friends drive | 0 (0%) |
| Transport service | 0 (0%) |
| Public transport | 0 (0%) |
| Occupation Status |  |
| Work, Full time | 4 (30.8%) |
| Work, Part time | 0 (0%) |
| Homemaker | 0 (0%) |
| Retired | 9 (69.2%) |
| Volunteer | 0 (0%) |
| Unemployed/seeking employment | 0 (0%) |
| Disabled | 0 (0%) |

### Thematic Analysis

The recording of the DTog COVID-19 Exit Questionnaire includes 333.36 minutes of responses and 702 total coding software references across all the coded subthemes. The transcribed responses of each DTog participant were coded in a range from 19 to 43 subthemes and categories.

DTog caregiver participants (n=13) revealed themes gained from the Exit interviews, which demonstrated caregivers’ thoughts and actions about caregiving activities, their personal lives and satisfaction, and their care recipients’ state during COVID-19. Subthemes revealed in the DTog Exit Interview can be found under **Table 2**.

### Technology: An Imperfect Solution to Social Isolation During Caregiving

This theme focuses on caregivers’ responses to the act of caregiving during COVID-19.

While there were several restrictions on activities due to social isolation requirements of COVID-19, caregivers maintained—and even found improvement in—many activities via use and adaptation to technology. However, restrictions on social and caregiving support activities remained.

Participants faced difficulties in accessing necessities. C4 shared her frustrations of the increased behavioral changes related to COVID-19 precautions: “Many businesses are not open or not wanting face to face interaction. [If] they haven’t, then they require a mask, which is an additional […] thing to keep up with.” Access to everyday needs such as food was limited, as C2 and C6 experienced understaffing issues at their care facilities. C2 describes, “[we] had to eat out of Styrofoam [for] all our meals, because they shut down all the dining rooms.” C6 attributed staff shortage to the desire to not “work in an environment they felt at risk.” Healthcare access was restricted for many participants (C1, C2, C4, C9, C11, C17). C9 was unable to access emergency care when “[the care recipient] fell and she cut her chin…bleeding.” C11 experienced a lack of COVID testing, having “to go and sit in a car for four hours, as we never even got one.” C1 expressed that her care recipient’s symptoms and progressions in dementia could have been noticed and addressed sooner if there were no delays in accessing healthcare and more social engagement that allows other sets of eyes.

Activity outside of the home was greatly limited due to the pandemic. Whether it was going to restaurants, stores, or friends’ houses, C11 had to be careful anytime they left home, to prevent the possibility of contracting and spreading COVID-19 to her care recipient. Maintaining social networks, habits, and relationships with others, including the care recipient, was consequently challenged. C6’s care recipient’s condition “was so bad” that there “was no reaction on her part” when C6 visited through restricted and complex visiting rules. C10 did not even attempt to visit her care recipient because of how prohibitive it was. C13 compared her visit to her care recipient to a “prison,” especially with the “timed” visits:

> “C13: it was a very cruel experience…if I did [manage to meet him], I had to see him in a window or I had a porch […] so it was […] sort of dehumanizing…it was horrible because I had to wear all this gear and very artificial and he would get upset and I would get upset.”Participants cited struggles in communicating with the care recipient. C1’s care recipient had difficulties in hearing and reading facial expressions with masking. C2 wears hearing aids that cannot process low tones well; as a result, conversing with their care recipient was very difficult, especially as the care recipient’s vocal volume decreased.

However, many participants used and adapted to advanced technology to carry on daily caregiving duties and habits, finding improved convenience. Participant C11 mentioned that she signed up for a “meal delivery plan” that was “tremendously” helpful, as she no longer needed to leave to buy groceries or spend time cooking, which was similarly noted by C2. C3 and C11 noted that shopping was made easier with apps like “Instacart.” Nine participants (C1, C2, C4, C6, C9, C10, C11, C13, C21) shared positive experiences of utilizing telemedicine and healthcare online. C1 found it pleasing that all the documents and information are in one place online, which simplified caregiver work. C2 and C4 related to the convenience of not driving; by avoiding traffic and the time spent driving, care recipients could be prevented from being left alone when the caregiver was at an appointment, putting caregivers more at ease. Having needed to “track [doctors] down” via phone in the past, C1 was a big fan of how doctors now responded “within 24 hours” through the patient portal. In fact, C11 referred to telemedicine as “the way in the future” and C21 looked forward to a future hybrid version of healthcare.

Social activities were maintained via technology. C1 “learned how to do Zoom” and C21 no longer felt scared of Zoom. C4 found the usage of virtual settings helpful for remaining in contact with “coworkers” and holding “weekly family calls.” Five participants (C2, C3, C4, C9, C19) mentioned that the assistance they received from family and friends has “alleviated [their] burden.” The interactions between care recipients and their social circles allowed the caregiver to perform other caregiving duties. Some participants, meanwhile, viewed the limited levels of socialization during the pandemic as a change for the better in caregiving. For instance, C1 became more “accustomed” to a “quieter lifestyle,” viewing COVID-19 as a “good teacher” that prepared one “for the changes the diseases [will] bring.”

Nonetheless, participants also reported on limited effects of technological aids, frequently revisiting the issue of restricted face-to-face interactions. Online social platforms did not alleviate the social loss for C21, who rather felt “stuck in the house a lot” and that “it makes things a little tougher emotionally.” Meanwhile, four participants (C3, C9, C17, C21) expressed their struggle due to the decrease in social interactions with others.

Several participants spoke on the insufficiency of virtual healthcare services. C6 and C9 felt that virtual appointments are ineffective because the physicians do not see how patients sit and move; C4 found that virtual appointments “lack the relational component”; C19 thought the duration of physician-patient direct interaction to be low, and remarked that most of the care was handled by nurses.

C4 expressed that healthcare service quality after COVID-19 had declined, saying that healthcare professionals were “not really cognizant of the fact they’re not providing great care or that they don’t care that they’re not providing great care.” C4 further pointed out communication challenges in telemedicine procedures:

> “[Businesses and doctor’s offices] require a mask, which is an additional […] thing to keep up with. And […] we’ve had where we showed up to a meeting that they’re like, oh, it’s virtual, we forgot to tell you, we sent you a letter in the mail, but, but the letter arrived, like three days after the appointment.”Our interviews show that the sudden shift to technology had become a barrier to some caregivers-especially with regards to continued participation in support groups. C19’s support group had not resumed at the time of the interview which was still during the pandemic. C6 shared: “The Piedmont [hospital] is running a simultaneous in person [and] virtual [support group] and they’re struggling with the technology. The [support group is] just not up with the technology yet.” C11 struggled to find online support groups specifically for LBD. Meanwhile, C2, C3, and C6 mentioned their struggles in finding in-person support services at care facilities and accessing additional resources for their caregiving.

### Self-Care is A Necessity Even During A Crisis

Self-reported experiences of caregivers show deterioration of emotional, physical, and social well-being to the extent of burnout due to the extensive nature of caregiving during the pandemic; however, coping methods involving social support, self-focused activities, and positive mindsets were found by caregivers, demonstrating resilience.

The sacrificial nature of caregiving activities contributed to caregivers’ deterioration of physical and mental wellbeing. C1 reported a decrease in exercise due to the desire to not leave their care recipient alone. C10 and C21 had to avoid physically accessing healthcare during the pandemic, due to caregiving commitments and the dangers of exposure to COVID-19. Despite taking safety precautions, like attending one-on-one appointments and masking, in-person visits were viewed as risky. The deteriorating conditions of his care recipient caused stress for C2, exacerbated by the pandemic situation to the point where he considered going to counseling.

“C2: I get pretty far down sometimes. How long can I keep doing this? It’s hard to keep [caregiving] every day, and to see your loved ones suffering […] is terrible to watch her (the care recipient) shriveling up.”

However, caregiving during a pandemic made caregivers persevere regardless of their health status, causing risk of mental and emotional burnout. C4 commented that they “have no work life balance” due to the extended responsibilities of COVID-19 caregiving. In fact, 3 participants (C10, C11, C13) cited the lack of self-care during caregiving, accompanied by the feeling of isolation due to the lack of solidarity and others’ understanding of LBD caregiving. C10 thought she was doing a good job as a caregiver until she “saw some friends in person” and realized she felt exhausted, struggled to communicate and explain LBD caregiving to others, and had no time for self-care like exercise and connecting with friends. C19 and C17 felt anxious about contracting and transmitting COVID-19 to their care recipients, “and not knowing how [COVID-19] was going to come down.” C11 struggled the most with the lack of knowledge and direct support in the caregiving field, which made caregiving more difficult and took away opportunities for independence.

Some caregivers expected greater stress and increased responsibilities in their caregiving future. C2 called COVID-19 a “vicious virus” and doubted the possibility of going “back to normal” in his “lifetime, which is not that much longer.” C1 believed caregiving “is gonna just get more challenging,” such as the overwhelming changes in responsibilities and restrictions, that put C1 to a state of misery. C11 had a hard time embracing accelerated divides of the nation due to differing beliefs, which made her reluctant to share her opinions. Some caregivers were doubtful of the quality of care for their care recipients and receiving caregiving support from healthcare workers. C3 found hiring other caregivers to be unreliable due to their inconsistencies with caregiving scheduling, making C3’s life “a lot more harder.” Despite the safety precautions, C13’s care recipient contracted the disease within her facility, which made C13 “very frightened.” C10 felt “infuriated” when seeing some workers at the care community who “either weren’t wearing a mask or weren’t wearing the mask correctly.”

A large component of the COVID-induced changes in caregiver habits and routines was from social activity, or the lack thereof. Multiple caregivers cited the decrease in social interactions made caregiving difficult, affecting their personal health. Five participants (C1, C2, C3, C4, C6) listed negative effects of reduced socialization with friends and family. C1 felt like her caregiving support was limited, while C4 found “intimate relationships” in both work and friend settings difficult to maintain—a drawback that was “tough mentally” for C2. Both C21 and C10 became aware of the importance and lack of physical connection with friends. C10 also realized a “rift” developed between him and his friends during COVID-19 due to differences in masking beliefs. When C17’s sister passed away, C17 was unable to attend the funeral due to feeling “reluctant to get on a plane” with the pandemic.

Nonetheless, caregivers found support systems and methods that helped them develop resilience to combat these burdens. As mentioned in the first theme, technology played a large role in improving access and convenience in caregiving, which further benefitted caregivers’ personal life. Several caregivers specifically found Zoom to improve convenience in their daily activities such as working at home (C1), accessing healthcare more conveniently (C1, C2, C4), attending support groups (C11), and staying in touch with friends (C1), which helped better manage work-life-caregiver responsibility balance (C1, C5).

Despite health guidelines and social distancing, caregivers found ways to engage in in-person social activities, on top of online interactions, that further helped alleviate care burden. C4 found comfort in locally knowing someone who understands and relates to her caregiving experiences and felt less socially isolated after moving closer to her extended family, friends living in the same neighborhood, and a small church group. Familial support helped reduce caregiver burden for five participants (C4, C2, C1, C3, C19). C4 and C3 received caregiving help from their sons to allow more time for themselves––such as attending physical therapy or having sick days off. C21 described the importance of family to her wellbeing:

> “C21: We just love [our kids and grandkids] too much to have them excluded from our lives, and they’re taking care and we’re taking care [when interacting in-person]. So it’s like you […] got to balance all the risk with the reward, and you need the reward so badly in your life, especially now we’re getting into a time of our life when you really know there might be some bad stuff coming down the road. You don’t want to waste anything.”Friends were also avenues of social support for caregivers. C10 attended in-person gatherings for walks and touched base on Zoom with their friends, feeling socially connected. For C3, their friend even helped look after her care recipient for three months, not only alleviating the caregiver burden but also developing a bond with the care recipient.

Successful examples of support expanded to external programs: C3 hired another caregiver who has “been able to handle all” in coming and giving the care recipient their medications, which provided C3 with downtime. C5 and C11 referred to the DREAMing Together program, specifically being able to interact and connect with their coach:

> C11: it was really interesting meeting someone who’s in med school now and going through […] not only […] COVID but with all the other crazy stuff that’s going on out there and just, […] what she’s learning, what she’s not learning, […] I really found […] her to be a great not only resource, but just I really enjoyed her a lot.”Caregivers utilized the COVID-19 challenges as opportunities for self-growth via developing new skills or reviving interests, showing traits of resilience. C1 was able to find time to self-reflect, develop new habits, and engage in new hobbies when staying at home during COVID-19, learn language and cooking online, and start independent and collaborative art projects with the care recipient. C6 had more time to catch up on projects and tasks after “putting [his care recipient] in a nursing home,” post pandemic. C10 revived her online store and found an outlet where she could put her energy and thoughts in, where she would “stretch in ways that I didn’t really know I could or expect.” C4 had the opportunity to refine her “telework and webinar facilitation skills” and earn a certification in “Project Management Professional.” On a similar note, C17 and C21 stated that they have acquired technological proficiency, such as utilizing Zoom, because of COVID-19.

Caregivers also developed positive attitudes to life, caregiving, and future plans despite burden as a caregiver during COVID-19, demonstrating resilience and adaptation. C2 and C11 learned to keep moving forward and not let unexpected circumstances prevent one from living their life. C4 found reliance and faith in God as important. C10 initially struggled to accept the changes to her living habits due to COVID-19 and her autoimmune disease, but eventually accepted that she will have to continue masking for her own safety. While C11 emphasized the continued importance of hygiene to prevent contracting COVID-19, C17 and C21 noted the importance of social interactions during COVID-19—where C21 learned to value the balance between personal safety and family time. Via the DREAMing Together modules, C3 learned how to ask for help from others, regulate anger, and become open-minded. In the perspective of a caregiver of older age, C13 acknowledged that it “becomes a lot more challenging [to let] go of guilt” of not being able to personally care for the care recipient and instead putting them in a care facility, but realized that this feeling of guilt is “a typical struggle that people have as caregivers” and that caregiving is not only caring for the care recipient, but also “about caregiving for yourself.” C21 expressed gratitude towards her caregiving experience, for it provided “purpose into [her] older age” and help “getting more patient.”

> “C21: So adapt or die […] and that’s my job these days, is to be quick enough at noticing what needs to be adapted to do it [on] time. And to do it gracefully [and] patiently.”

### Caregivers Report A Reinforcing Negative Cycle of Declining Care Recipient Health and Adaptability

This theme demonstrates caregiver observation and description of how COVID-19 impacted care recipients. Despite only one question asking caregivers to share directly on their care recipient’s state, caregivers ended up sharing a lot about their care recipients. It is important to note what caregivers notice in their care recipient as it reflects their mindsets on caregiving, especially after going through the caregiving resilience coaching program, and the care recipient’s condition both influences and reflects caregiving directly.

Caregivers identify a reinforcing negative cycle imposed on care recipients by COVID-19: Social isolation restricted social activities and communication, which accelerated the already-present decline in physical, mental, and emotional health from LBD; this limited the care recipient’s ability to adapt and be resilient, further exacerbating their condition (See **Figure 1**).

**Figure 1.**
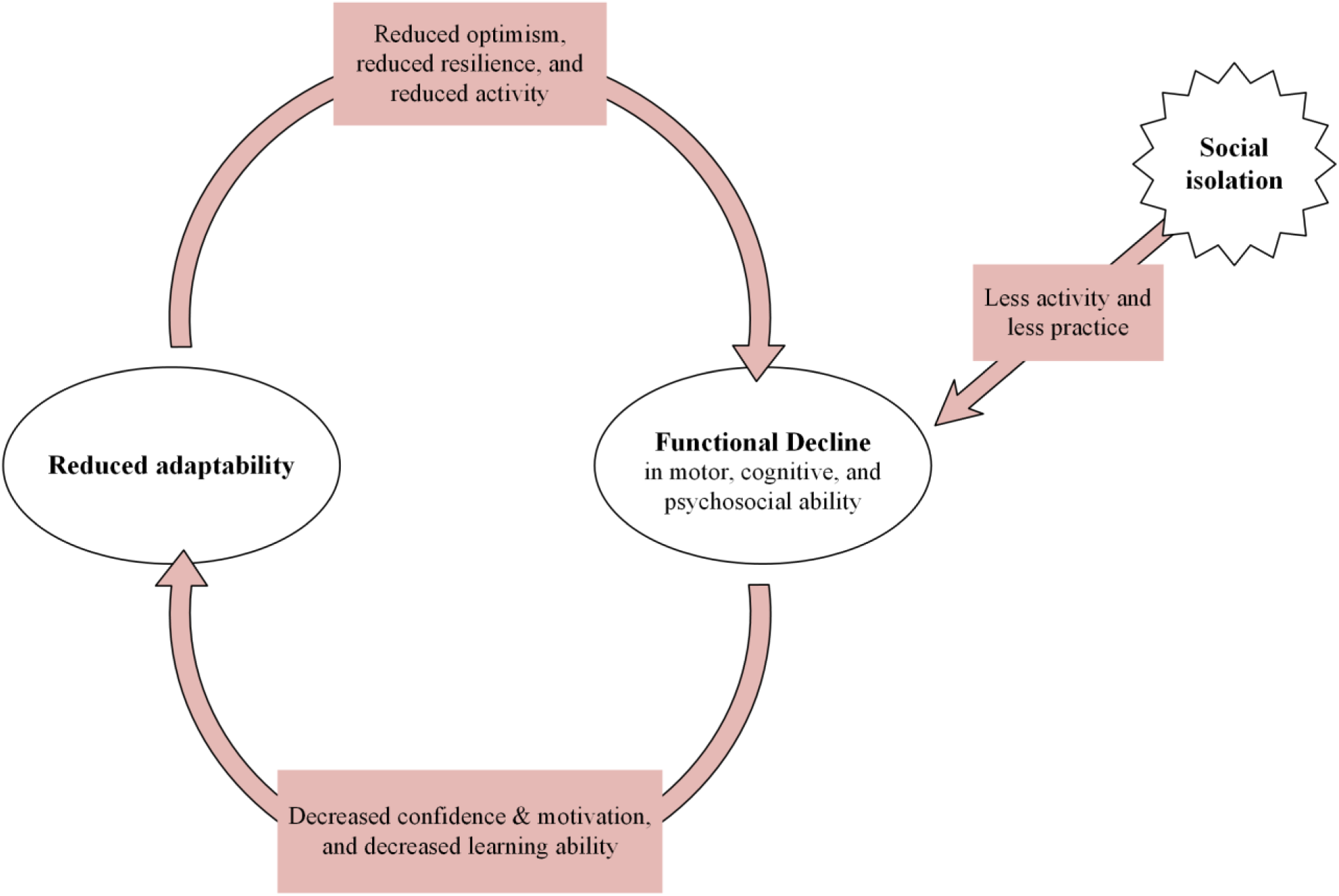
Diagram demonstrating third theme: Caregivers Report A Reinforcing Negative Cycle of Declining Care Recipient Health and Adaptability

While communication was already a difficult skill for people with LBD due to motor-cognitive issues, the pandemic caused further limitations as social gatherings were discouraged. C1 reported that her care recipient struggled with reading expressions and hearing others due to masking, such as in doctor’s appointments. C3’s care recipient shared this struggle and started speaking less as a result. C19 added, “[My care recipient] just doesn’t want to talk […] because people want to ask him questions. And psychologically he feels he is not able to answer them.”

While caregivers were able to adapt to technological dependency in all aspects of COVID-19 life, they report that their care recipients remained struggling with technological advancements, attributed to their LBD-invoked motor-cognitive decline. C1’s care recipient consistently needed assistance in using Zoom, while C19’s care recipient could not and would not use Zoom despite the assistance of nurses. Caregivers reported that Zoom confused (C9) and “frightened” (C13) their care recipients.

Caregivers noticed further decline in care recipient cognitive health, such as recognition, cognitive processing (C16), and concentration (C3), after pandemic-onset. C2 found their care recipient “struggle” to correctly answer questions on mental tests, believing that the lack of exercise exacerbated the deterioration. C5 also thought that the decrease in activity is what “contributed to him [care recipient] slowing down [his cognition],” as he “felt isolated because of the pandemic.” Four other caregivers attributed the cognitive deterioration of their care recipient to reduced socialization (C1, C2, C9, C11, C21). C2 and C11 described how their care recipients were inflicted by hallucinations and cognitive decline, which was exacerbated due to social isolation:

> “C11: He thought somebody was breaking into the house, and he ended up calling the police. […] He was hearing things and saying things, […] he just didn’t do well on his own. […] I think it was […] being cooped up in a house all day […] and not having the social interaction that he was used to having.”Emotional health was impacted by social isolation as well: C10 reported that his care recipient did not have many people to talk to at her care facility, thus struggled with “making friendships” and, consequently, loneliness. C17 attributed her care recipient’s depression to the lack of socialization, especially as someone who “likes to socialize a lot.” The recipient also maintained fear towards COVID-19 despite living in the pandemic for “over two years,” which C17 believed played a role in this emotional decline. C11 and C10’s care recipients shared this fear and caused C10’s care recipient to relocate to a care facility closer to C10. Further, C13’s care recipient experienced paranoia when he was moved to a care facility during COVID-19, because he was unable to process changes due to the pandemic:

> “C13: He [care recipient] knew […] I would say this is all because of COVID rules, […] that I can’t bring in the virus [while visiting him at the care facility], and they’re [the care facility is] trying to take care of you. […] But he was agitated. He couldn’t totally process it like you or I could, […] so […] he was frustrated with it.”Along with a rapid impairment of functional ability, C13’s care recipient also felt a lost sense of purpose due to COVID-19 shutting down festivals and businesses, which had “gave him a great purpose in the morning and go do his paintings” in the past.

Physical health decline could not avoid this exacerbation cycle either. C6 and C11’s care recipients “continued to lose weight” due to difficulties accessing food while having to live independently apart from the caregiver. C19 noticed increased tremor in their care recipient living in an assisted living facility due to delayed delivery of medication, especially in the 2021 COVID year. Caregivers reported on decreased physical activity in care recipients, as C2’s care recipient’s “group fitness classes…were no longer an option after the pandemic,” and C9’s care recipient “was much more mobile [before COVID-19] than she is now [during COVID-19],” which led both to being less active. C4‘s care recipient directly contracted COVID-19, which resulted in greater physical imbalance.

A few caregivers shared instances of their care recipients maintaining cognitive and emotional health, specifically via finding ways of socialization for emotional support: “A lot of communication and keeping in touch” with family supported C2’s care recipient as it “helped mentally”; C21’s “sweet” attitude helped their care recipient improve emotionally and reduce stress; C3’s interaction as a caregiver helped her caregiver become “much more lucid” and decrease hallucinations. On the other hand, C1 shared that her care recipient was unable to “get involved in a conversation because there’s too much going on” due to the progression of his LBD, but found the decrease in social interactions to be beneficial for taking “some of the pressure off” and getting “accustomed […] to a quieter lifestyle, sort of less social interaction, without it being […] sudden.”

## Discussion

The participants in our study—LBD caregivers that completed a caregiving resilience coaching program—fulfill our hypothesis that activity helps grow resilience. Despite the heightened risk of mental and emotional burnout from exacerbated caregiving burden from pandemic circumstances, DTog participants were able to avoid so in two ways: 1) engaging in meaningful activities to prevent feeling overwhelmed by caregiving burdens, and 2) establishing positive mindsets and beliefs to adjust to new caregiving responsibilities from the COVID-19 pandemic. Caregivers demonstrated resilience by overcoming obstacles and re-establishing activity equilibrium, which resulted in improved life satisfaction. This aligns with the Activity Theory of Aging in that maintaining a high level of activity relates to increased life satisfaction and well-being.

While successful coping strategies involved activities, a commonality was that these activities were unrelated to caregiving and allowed caregivers to live a role other than a caregiver. Those who went a step further and utilized the COVID-19 circumstances as an opportunity—a stronger sign of resilience—did so by participating in self-caring and self-motivated activities, suggesting the importance of self-care and preventing caregiving from over-dominating one’s activity.

In fact, our research team conducted a similar study involving a Focus Group who did not participate in the program in 2020, earlier in the COVID-19 pandemic (Chan et al., 2025). While the caregivers in the Focus Group were mainly concerned about caregiving roles and were still struggling with social isolation, DTog caregivers in this study introduced and highlighted the importance of self-care, already having implemented successful strategies around restrictions.

Though it may be that these participants demonstrated greater adaptability because more time has passed since the onset of the pandemic, it is also a possibility that there is a relationship between self-care and resilience as an LBD caregiver during difficult times.

Hence, the rationale that equilibrium is essential to being a healthy caregiver can be presented. The role accumulation theory establishes that fulfilling multiple social roles benefits one’s psychosocial well-being (Sieber, 1974), yet the overwhelming nature of caregiving is prohibiting this benefit. Recent research specifies that roles need to be voluntary, rather than obligatory, for beneficial effects in older adults (Mize et al., 2025). As most caregivers were older adults and loved or related ones to their care recipients, they may have started caregiving without much choice, planning, or compensation; therefore, it is likely that caregiving was considered obligatory work at least partially, causing mental burden instead of benefits to psychosocial well-being. This may explain how the self-motivated coping activities included independent, solitary activities, which seemingly contradicts the Activity theory. This is a recurring idea from our previous research with the Focus Group (Chan et al., 2025), which also suggested that anything excessive is harmful. Balance of identities may be needed for pleasurable activity. It could also be considered that the Activity theory is based on the need to maintain the same level of social activity as they had when they were younger; for people with lower base levels of activity, caregiving may exceed that level and cause exhaustion.

It is important to note that the successful coping activities of caregivers served a purpose more than just a distraction from caregiving, as caregivers also felt supported while bonding over caregiving struggles. With balance maintained, caregiving could become part of self-development and identity to positively motivate the caregiver, such as how DTog participants built positive mindsets. Another rationale is also possible: LBD caregiving is burdensome due to the lack of emotional fulfillment. Bonding over caregiver struggles did not engage the caregiver in a different social role but still provided a relieving effect, suggesting that emotional support is a key necessity to caregiving. Recent studies highlight loneliness as a major contributor to caregiver burden. Emotional loneliness was associated with caregiver burden for care recipients dealing with dementia and mental illnesses. This result was not consistent for caregivers of people with solely physical illnesses (Hajek et al., 2021), suggesting that cognition may play a bigger role in emotional engagement and that caregiving especially needs support that is emotional. In fact, unpaid caregiving itself is a factor associated with higher levels of loneliness (Grycuk et al., 2022), further highlighting loneliness as an important factor. In our study, the emotional aids—such as comfort, connection, and enjoyment—as well as the lack thereof, were consistently mentioned when caregivers discussed support and struggles that they have experienced, respectively. A recurring theme for both the caregiver and care recipeint was difficulty in communication: Even when the care dyad was presented with constant social interaction opportunities while being together during the pandemic, communication barriers restricted emotional engagement. Thus, it may be that the Activity theory stands only when the activity involves *emotionally-fulfilling* activities. While social activities often play a role in fulfilling people emotionally, when a social role induces emotional burden, escaping the role and the burden—thus restoring positive emotional fulfillment—may be prioritized over maintaining social activities. In fact, research showed that, for caregivers of people with mental and physical issues like LBD, levels of burden, loneliness, and social isolation were significantly higher after the pandemic than before (Hajek et al., 2021). This supports the idea that COVID-19 exacerbated emotional burden in LBD caregivers.

Though technological systems did allow care dyads to continue some of their daily activities, including those related to caregiving, it was limited. Daily, necessary, and social activities were still heavily restricted, causing mental and physical stress. Participants desired *physical* social interactions, because virtual interactions lacked the feeling of connection. Thus, technological aids may not have been fully beneficial due to their insufficient socioemotional support. Future development of support systems for caregiving, therefore, should address this socioemotional necessity.

## Conclusion

The presence of the Activity Theory and activity as an element that helps build resilience was observed in LBD caregivers who participated in a caregiving resilience coaching program. Caregivers overcame COVID-19 and caregiving obstacles via an active attitude, which was associated with decreased burden and higher life satisfaction, and reflected adaptability and resilience. However, our findings suggest that caregivers specifically need emotionally fulfilling activity, perhaps that which preserves their identity other than being a caregiver, in order to alleviate the mental and physical burden. Future development of support systems for caregiving, therefore, should involve this socioemotional necessity.

This research has important implications for the development of LBD care support programs. It calls for more research and caregiving support programs like DTog for LBD caregivers, as this study is one of few that addresses LBD caregiving in general. This analysis informs solutions, including educational programs, therapy, and future studies, to address the multifactorial difficulties of LBD caregiving, the lasting impacts of the pandemic yet to remain for LBD care dyads, as well as upcoming obstacles to which new and old caregivers need to adapt to, with today’s ever-changing world.

## Limitations and Future Directions

The study has several limitations. The relatively small sample size is considered to limit the generalizability and merit of our conclusions. Women tend to express higher levels of loneliness than men, and older adults with less education tend to experience greater social isolation than their counterparts (Bay et al., 2020; Tanskanen & Anttila, 2016), which could influence responses. Due to the involvement of different interviewers, questions and probes may have been asked differently in the COVID-19 Exit Interview in meaningful ways. While efforts to mitigate acquiescence and response bias were made through a standardized question guide and open-ended question stems, these biases may still have occurred because of the different interviewers and their tone, presence, and rapport with the participants. Future studies will look at DTog program, with more participants and in its entirety, to identify more specific, beneficial effects of the program on LBD caregiving and its potential as a viable support solution.

## Data Availability

All data produced in the present study are available upon reasonable request to the authors

## Conflicts of Interest

The authors have no conflicts of interest to report.

## Data Availability

Data is available upon request and stored in a protected network.

## Funding

The study was funded by the Emory University Roybal Center for Caregiving Mastery with funding number P30AG064200, the Emory University Center for Health in Aging, and by the Parkinson Foundation.

## Acknowledgements

The authors would like to sincerely thank the caregivers and care recipients who generously shared their time, stories, and experiences with us. The authors are also grateful to Dr. Madeleine Hackney and her lab for their endless and unconditional guidance, feedback, and support.

Finally, the authors would like to thank Guy Harris, Cecelia Lofton, Emma Rose Brown, Emma Macmanus, Sara Popofsky, and Shawo Lhabdon for their support in the coding process.

## Supplemental Material

**Supplemental 1.** The DREAMing Together Program Intervention The DTog program is composed of weekly readings, recordings, and conversations for eight weeks. The program was developed iteratively by the investigative team, drawing from other patient facing resources, literature, and foundation materials for patients. Medical and graduate public health students, and clinical research staff acted as coaches to lead the calls and sessions. DTog coaching was conducted virtually. 8 weekly modules guided caregivers through coaching exercises and strategies intended to improve caregiver mastery. Topics included an Introduction to Caregiving; Understanding Dementia and Caregiving; Diagnosis of LBD, PDD, DLB, and others. (See **Table 4**). Within each weekly module, there were visual diagrams, activities, worksheets, personal stories and anecdotes, and informative readings regarding strategies to increase caregiving resilience, health disparities, how caregivers can address challenges with their care recipients, and maintain a healthy lifestyle. Each module was approximately 20-30 pages of reading, or 1.5 hours of audio to be inclusive of those with low literacy or who preferred an audio version. No participants needed the audio version.

**Table 4.** Interventional Pedagogy: Dreaming Together (DTog) Program 8 Weekly Modules.

|  |  |
| --- | --- |
| **Weekly Modules** | **Module Content Summary** |
| Week 1: Introduction to Strategies for Developing Caregiving Mastery for Those with Lewy Body Dementias | • Introduction to the DTog Program which includes an overview of what it means to be a caregiver and examples of well-known caregivers<br>• Discusses how to know when it is time to “step up” and begin providing more care for someone who has been mostly independent in the past |
| Week 2: Understanding Dementia Caregiving | • This module concerns caring for someone with Lewy Body Dementia and other related conditions that impact one’s memory<br>• Multiple ways to support someone experiencing memory conditions |
| Week 3: Diagnosis & Treatment of LBD, PDD, DLB, and Others with Similar Conditions | • The symptoms of these dementias and what is known about these conditions is described in this module<br>• Information on how to get involved in research on LBD is provided |
| Week 4: Increasing Public Awareness and Advocacy | • This module provides strategies for increasing public awareness of the challenges of caregiving and being an advocate for other caregivers and people with dementia or other memory problems |
| Week 5: Helping Your Care Recipient Sleep | • Helping the care recipient sleep, especially when that person is a spouse or life partner since sleep problems are common in LBD and can be difficult to manage<br>• This module will also go over methods to help someone with sleep problems—and even help caregivers get better sleep |
| Week 6: Self-Care and Prevention Interventions for Caregivers | • The module is about the caregiver health, teaching caregivers the importance of self-care and how and where to find support<br>• Provides tools and strategies to help caregivers remain healthy and relax while also being in the role of a caregiver<br>• Discuss how the type of relationship caregivers have with someone can make it harder to care for them, and give ways to manage difficult relationships |
| Week 7: End of Life Care | • Focuses on the care that one receives towards the end of their life, including various options to help caregivers and their care recipients be prepared for the future<br>• Options for end of life care and explores misconceptions<br>○ Includes advance care planning, palliative care vs. hospice, and spiritual and cultural preferences when it comes to care at the end of one’s life |
| Week 8: Program Recap and Hearing from the Ones in the Trenches | All sessions are reviewed and tied together. Strong focus on empowering caregivers by learning from the experiences of others in similar situations |

Every week, participants engaged in a call or online video conference. Most participants used the video conferencing application but often without the video option. Calls were conversations structured with an interview guide and were to last 30 minutes. Participants were probed regarding the modules to ensure participants developed a strong understanding of the lesson. Participants were asked how they were using the new strategies to improve their caregiving self-mastery in their daily lives. The calls engaged the participants with questions and topics that encouraged them to summarize and reflect on recent readings, relate, and apply the lessons to personal life, or share caregiving experiences. Questions were open-ended and structured to facilitate participation, minimize acquiescence bias, and allow participants to freely interpret the questions (**Table 5**). Participants were encouraged to discuss and share unique experiences related to the program topics. If necessary, gentle prompts brought the conversation back to the topic. Probes were used to obtain clarification and elaboration for all participant responses (**Table 5**). The theoretical basis of these processes is that the more a participant actively engages with the content, the more likely they will store the newly acquired information and incorporate it into future caregiving (Griffiths & Keirns, 2015; Havighurst, 1961; Zhaoyang 2021). Module coaching sessions ended with final thoughts and feedback on the modules and content. The DTog Program was built on the previous, successful DREAMS programs (Dillard et al., 2018; Hart et al., 2017; Perkins et al., 2019; Schindler et al., 2022; Shah et al., 2023b).

**Table 5.**
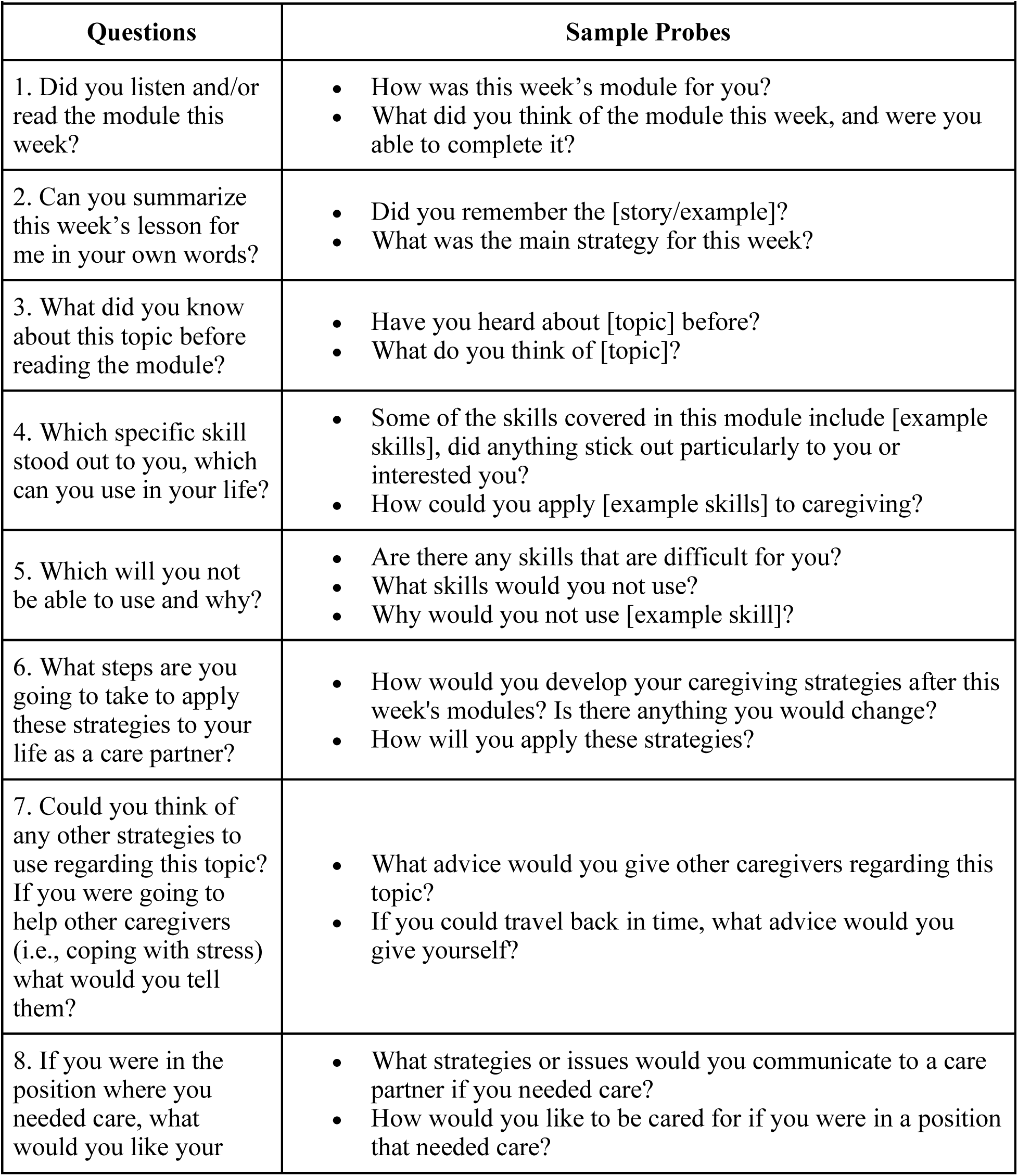
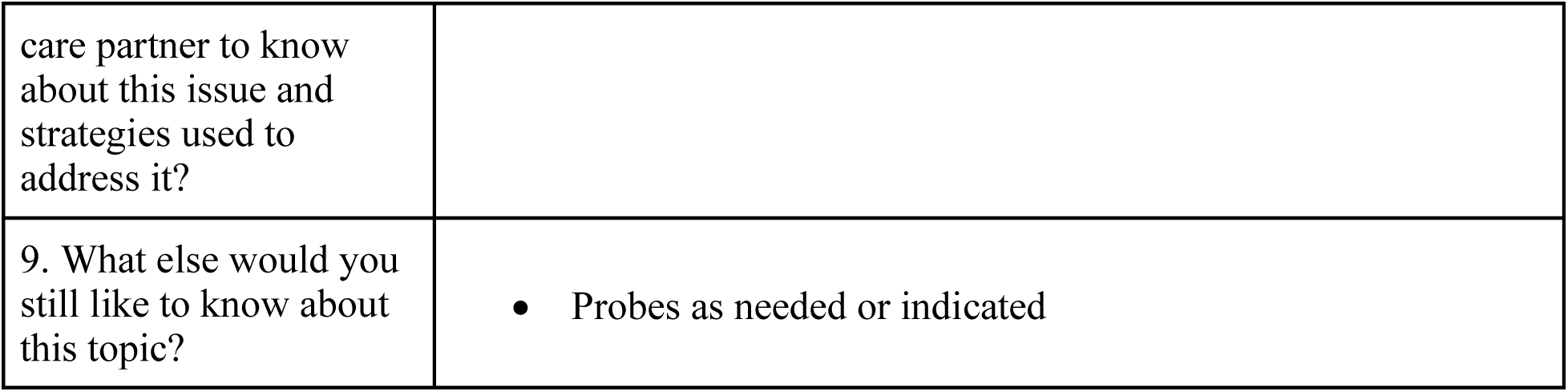
Interventional Pedagogy: DREAMing Together Caregiving Weekly Module Curricula Coaching Guide.

## Notes

### Competing Interest Statement

The authors have declared no competing interest.

### Funding Statement

The study was funded by the Emory University Roybal Center for Caregiving Mastery (P30AG064200), the Emory University Center for Health in Aging, and by the Parkinson Foundation

### Author Declarations

Institutional Review Board of Emory University gave ethical approval for this work (IRB00080676).

